# Altered EEG patterns co-vary with specific improvements in mind health at summer camp

**DOI:** 10.64898/2026.09.09.26362570

**Authors:** Narayan Puthanmadam Subramaniyam, Ryan Brown, Emma Gibbens, Anja Forche, Christopher Dirks, Jennifer Newson, Tara C. Thiagarajan

## Abstract

The mental health and wellbeing, or ‘mind health’, which we define as the ability to navigate challenges and function productively, of adolescents has declined substantially over the last two decades, corresponding with fundamental changes in their daily environments. These include the widespread adoption of smartphones and social media, as well as reduced sleep and time spent in nature. Device-free, nature-rich summer camps represent a substantial shift in environment and have been associated with behavioral and mental health benefits. However, the neural changes accompanying such environmental shifts remain poorly understood. Here, in a prospective pre–post study, we examined EEG spectral and complexity measures in adolescents (*N* = 30) before and after a three-week device-free, nature-rich residential summer camp. Following three weeks of camp participation, participants exhibited steeper aperiodic slopes, increased DFA scaling exponents, reduced entropy and Lempel–Ziv complexity, and decreased alpha and beta power, a convergent pattern consistent with a shift toward more structured or less random neural dynamics characterized by relatively greater inhibitory influence. These neural changes co-varied with reduced restlessness and hyperactivity, improved focus and sleep quality, and improved overall mind health in a subsample (*N* = 23) with both EEG recordings and self-reported mind health data. Precision analyses further indicated that participants exhibiting EEG changes beyond a defined threshold were also likely to show improvements in specific mind health metrics, with precision reaching up to 100%. Overall, participation in a device-free, nature-rich residential camp was associated with measurable shifts in neural dynamics alongside improvements in self-reported mind health, suggesting that sustained changes in the adolescent environment may influence both neural and psychological functioning.

**Trial registration:** ClinicalTrials.gov NCT07761143.

## Introduction

Adolescent wellbeing has been in sharp decline over the past two decades, concomitant with radical shifts in the environment [1, 2]. For example, unlike previous generations, the contemporary adolescent environment is defined by near-constant digital and social media exposure [3]. In the U.S, up to 95% of youth (ages 13-17) report using at least one social media platform, and over one-third report near-constant use [4]. Understanding what this environmental shift means for adolescent development requires asking not only what digital environments do, but what they displace — and what happens when they are removed.

Higher levels of digital and social media use are associated with lower self-esteem [5], reduced wellbeing [6], depression [7–9], anxiety [5, 10, 11], loneliness [10], and disrupted sleep [12–15]. At the same time, social media platforms can also provide opportunities for connection and support, with studies reporting associations with greater perceived social support, improved quality of life, improved well-being, and reduced stress [16–18], while educational literature also suggests many positive ways in which smartphones and internet can contribute to learning [19]. Nevertheless, the aggregate portrait of saturated digital environments is one in which attentional demands are high, restorative experiences are limited, and nature exposure is reduced.

Conversely, both nature exposure and in-person social interaction represent dimensions of daily life that digital environments tend to displace, and each has been independently linked to improvements in adolescent wellbeing [20–24].

Device-free residential summer camps introduce an environmental contrast by temporarily removing digital media and replacing it with outdoor activities and face-to-face social interaction. Nature exposure engages restorative processes as described by Attention Restoration and Stress Recovery theories [25, 26], while social belonging and peer connectedness are independently strong predictors of adolescent wellbeing [22, 23, 27]. Furthermore, in-person social interaction, but not social media interaction, has been shown to specifically predict adolescent wellbeing [24], suggesting that the structured removal of digital mediation may create conditions particularly conducive to the kind of meaningful peer relationships that screen-based environments tend to displace. Limited research has documented behavioral and socio-emotional benefits associated with participation in such camps, including improvements in self-esteem, confidence, independence, social cognition, and peer relationships [19, 28–31]. Using a larger sample of adolescents (*N* = 192) attending device-free residential camps, [32] found significant improvements in self-reported mind health and social belonging following camp participation.

However, these studies have relied almost exclusively on self-report and behavioral measures. Whether the environmental shift from a smartphone and social media-saturated daily life to a device-free, nature-rich setting produces measurable changes in brain dynamics and how any such changes relate to mental health outcomes remain entirely unexamined. Characterizing neurophysiological changes that arise concomitantly with behavioral and socio-emotional change would provide insight into the neural mechanisms through which device-free, nature rich environments influence adolescent development.

EEG provides a cost-effective way to characterize the neural correlates of both environmental and social influences on wellbeing. With respect to nature exposure, EEG studies have reported increases in alpha/theta [33, 34] and decrease in beta [35, 36] during nature exposure, while urban environments are associated with higher beta and gamma activity, associated with alertness and stress [34, 37]. Studies employing fNIRS have also shown that passive exposure to natural environments reduces prefrontal cortex activation, reflecting decreased cognitive load and enhanced psychological restoration [38, 39]. In contrast to nature exposure, the resting-state neural correlates of sustained in-person social engagement and belonging in adolescents remain largely unexplored, representing a significant gap given the documented importance of peer connectedness for adolescent wellbeing [22, 23]. However, much of the existing neuroimaging research has relied on brief laboratory exposures to pictorial, video, or virtual representations of nature rather than sustained real-world experiences. In addition, most studies have been conducted in adult samples and have rarely integrated neural measurements with comprehensive behavioral assessments of well-being. The neural mechanisms through which prolonged, naturalistic environmental interventions that combine nature exposure and social interaction influence behavior and socio-emotional status remain poorly understood. In this context, device-free residential summer camps provide a rare opportunity to examine whether sustained exposure to nature-rich, socially immersive environments produces coordinated changes in both self-reported well-being measures and neural dynamics.

EEG is particularly well-suited to address this question in naturalistic settings, given its low cost, portability, and temporal resolution relative to fMRI or fNIRS [40, 41]. Here we look at changes in two broad aspects of EEG signals before and after camp.

First, conventional spectral power analyses have been widely studied and shown to change with stress, with alpha power consistently decreasing and beta power increasing under psychosocial stress conditions [42], as well as varying systematically across a range of psychiatric and mental health conditions [43]. Second, we examine complementary measures of neural signal complexity, which characterize how much organized structure is present in EEG signals. A purely random signal like white noise has no temporal structure, whereas a highly organized signal has predictable patterns. Healthy brain dynamics occupy the territory between these extremes, and measures such as Lempel-Ziv complexity, sample entropy, spectral entropy, the DFA scaling exponent, and the aperiodic 1/f slope each quantify different aspects of this structure, with more random signals producing higher entropy and complexity values, flatter 1/f slopes, and lower DFA exponents. Alterations across these measures have been reported in several disorders. For example, elevated LZC has been reported in schizophrenia and depression [44], altered DFA scaling exponents have been found in major depressive disorder [45], and differences in the aperiodic 1/f slope have been identified in children and adolescents with ADHD [46], together suggesting that these measures capture fundamental properties of brain organization that spectral power analysis alone do not reveal.

In this study, we examined EEG recordings along with self-reported changes in mind health in adolescents attending Maine Teen Camp, a device-free residential summer camp in Maine that emphasizes nature engagement and in-person social interaction. We use the term mind health to reflect the broad, functional spectrum of capacities and symptoms captured by the mental health quotient (MHQ), distinguishing it from the common tendency to equate mental health with emotional distress and wellbeing with happiness. Assessments of mind health together with EEG recordings were carried out on both the first and last day of the three week camp. By integrating mind health and neural measures in a real-world adolescent sample, this study aims to determine whether the improvements in mind health associated with device-free camp participation are accompanied by shifts in neural dynamics. As a pilot study, we aim to establish the feasibility of detecting measurable EEG changes associated with an environmental shift, and to characterize the nature of those changes and their relationship to self-reported mind health.

## Materials and methods

### Study registration

The present EEG study was conducted as part of a broader prospective pre–post study examining changes in adolescent mind health and wellbeing during attendance at Maine Teen Camp (MTC), the behavioral findings of which have been reported previously [32]. The study was retrospectively registered at ClinicalTrials.gov (NCT07761143; https://clinicaltrials.gov/study/NCT07761143). Participant recruitment and follow-up were conducted between July 1, 2024 and August 17, 2025. The study was initially conducted as a prospective pre–post study of a naturalistic summer camp environment and was not considered a clinical trial at the time of participant enrollment. The authors confirm that all ongoing and related trials for this intervention are registered.

The trial registration encompasses both mental health outcomes and pre–post EEG measurements. The present study reports analyses of the EEG component and its relationship to changes in mind health.

### Camp environment

Maine Teen Camp (MTC) is an American Camp Association-accredited residential summer camp in Porter, Maine, USA, for adolescents aged 12–17 years [32]. During camp, personal phones and other internet-connected devices are prohibited and are surrendered to the camp office. Campers spend substantial time outdoors and participate in recreational activities and in-person social interaction with peers. The camp operated according to its usual program during the study, and the research team did not otherwise control the camp environment [32].

### Data acquisition

Participants were recruited from adolescents aged 12–17 years attending MTC, a device-free residential summer camp, during the periods from mid-July to mid-August in 2024 and 2025. As part of the broader study, 480 campers were invited to participate, of whom 333 began the assessment and 192 completed both pre- and post-camp assessments [32]. Campers were invited to participate in the EEG study that included electroencephalography (EEG) recordings at the start and end of the three-week camp period along with a comprehensive mental health assessment.

The final sample consisted of 30 adolescents where 23 attended MTC in 2024 and 7 attended the camp in 2025. Sample size was constrained by the number of EEG recordings that the team was able to collect within a single day. No a priori power-based sample size calculation was performed for the EEG component of the study. The EEG data were pooled across cohorts for analysis. The EEG data were pooled across cohorts for analysis. Parents received an information sheet describing the study prior to arrival at camp. Both parental consent and adolescent assent were required for participation. Ethical approval was granted by the Health Media Lab Institutional Review Board (HML IRB; OHRP Institutional Review Board number 00001211; Federal Wide Assurance number 00001102; IORG number 0000850). All procedures were carried out according to the Declaration of Helsinki. Participant flow through the broader study and the EEG component is summarized in Fig 1.

**Fig 1.**
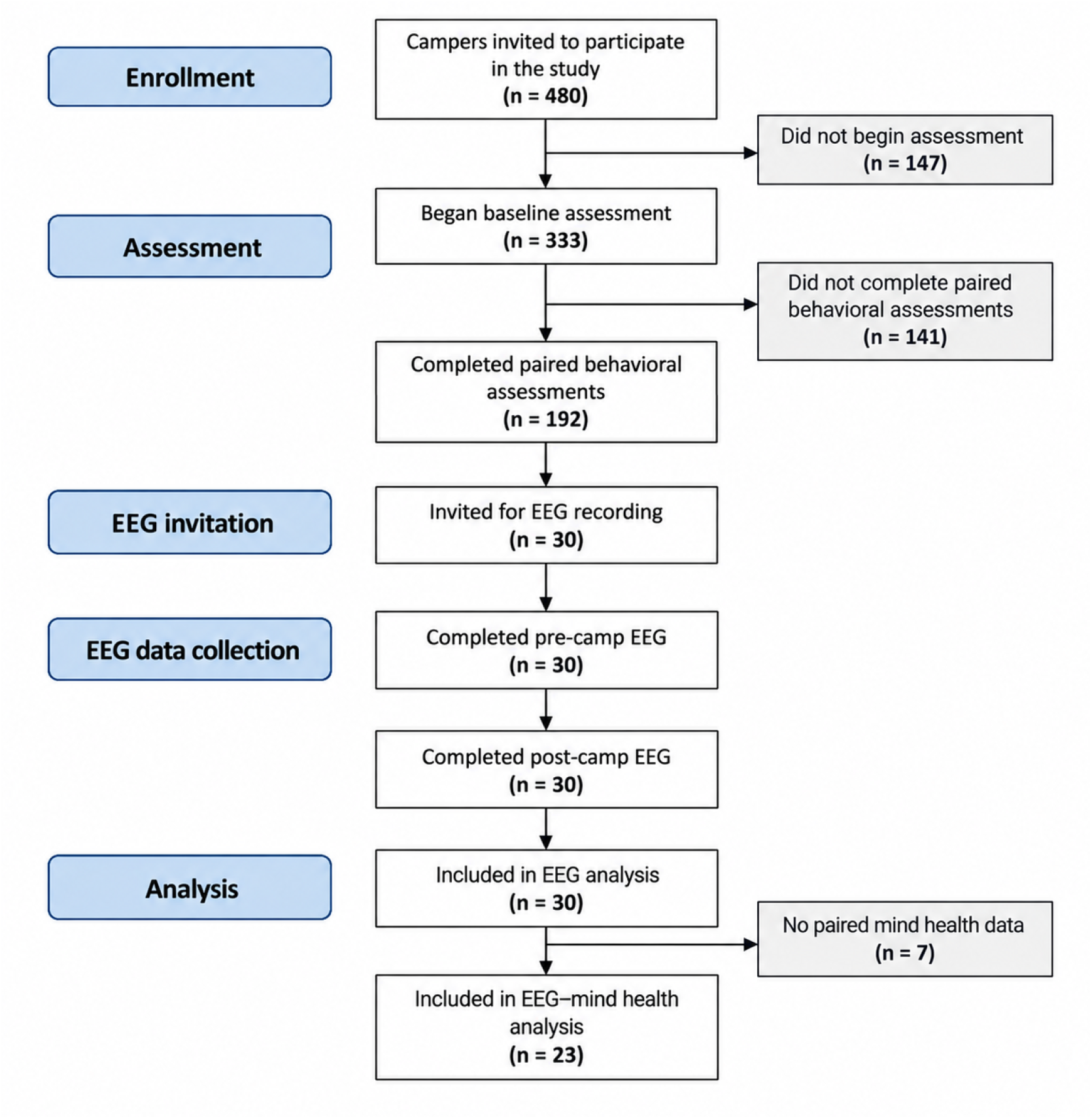
Participant flow through the study. Flow of participants from enrollment in the broader prospective pre–post study through behavioral assessment, EEG data collection, and analysis.

EEG recordings were acquired on the day of arrival at camp and again after three weeks on the day before departure from camp using an Emotiv FLEX2 Gel EEG system. Sixteen electrodes were selected from a standard 32-channel montage and positioned according to the international 10–20 electrode placement system. Mental health was evaluated using a 47-item Mental Health Quotient (MHQ) assessment, adapted from the adult version to ensure age-appropriate language and reading level; this youth-adapted MHQ is currently used in the Youth Global Mind Study (see Supporting Information for the full questionnaire). The MHQ queries 27 mental capabilities and 20 problems on a 9-point life impact scale. The capacities scale is bi-directional where 1=causes me difficulties and 9= is an asset to my life while 5 is neutral, while the problems scale is unidirectional with 1=Not a problem to my life and 9=has a severe impact on my ability to function. These 47 items span all symptoms queried across 126 different psychiatric, neuroscientific and psychology assessments used in research and clinical practice and include numerous aspects such as sleep quality, sadness and hopelessness, anxiety, restlessness and hyperactivity, stability and calmness, emotional control, anger and numerous other factors potentially associated with environmental factors such as exposure to nature and removal of devices.

### EEG recordings and preprocessing

EEG was collected while the participant was sitting quietly with their eyes closed using the EMOTIV FLEX Gel system. Data was obtained from 14 channels as shown in Fig 2 (AF3, F7, F3, C3, FC5, P7, O1, O2, P8, FC6, C4, F4, F8, AF4). The signal was sampled at 256 Hz and band-pass filtered between 0.5 and 40 Hz. All the preprocessing of the EEG data was done using the MNE python package [47]. Bad segments from EEG data were detected and removed using the MNE-auto reject algorithm [48].

**Fig 2.**
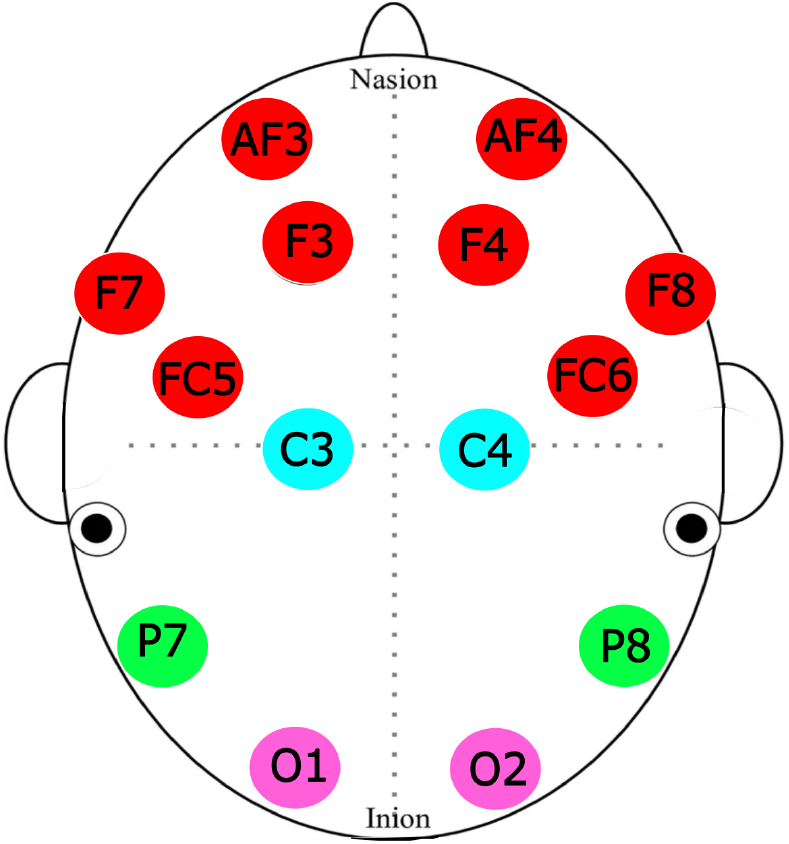
Location of 14 channels used on the EMOTIV FLEX Gel system. The channels were divided into Frontal (red), Central (cyan), Parietal (green) and Occipital (magenta) regions.

### Feature extraction

The following features were extracted from the data, after segmenting the data into 5 second windows.

#### Spectral power

The power spectral density (PSD) was estimated using the Welch method, with a 1-second Hamming window and 50% overlap between successive windows. Relative band power was calculated for delta (1.0–4.0 Hz), theta (4.0–8.0 Hz), alpha (8.0–13.0 Hz) and beta (13.0–30.0 Hz) frequency ranges.

#### Aperiodic slope

The aperiodic slope of the EEG power spectrum was obtained using the Fitting Oscillations and One-Over-F (specparam) algorithm [49]. The specparam algorithm models the aperiodic slope and then fits the oscillatory peaks with Gaussians. Gaussians are then iteratively subtracted until all the peaks are removed. Using an exponential function in the semi-log power space, aperiodic slope of the power spectrum is refit with the peaks removed, giving the estimates for the slope and the offset. Model fitting was performed on power spectra in the 2.0–30.0 Hz range, with the maximum number of oscillatory peaks set to five and peak width limits constrained between 0.5 and 12 Hz. The model was subtracted from the full power spectrum to obtain the aperiodic slope, which reflects power law (1/f behavior) and has been shown to be related to the changes in excitatory-to-inhibitory balance (E/I balance) [50].

#### Lempel-Ziv complexity

Lempel-Ziv complexity (LZC) is a metric to estimate the complexity of a discrete-time signal [51]. LZC analysis is based on coarse-graining of signals [52]. When applied to an EEG signal this involves transforming the signal to a finite symbol string *P*, for e.g. a binary sequence of zeros and ones based on a threshold defined as the signal median [53]. The string *P* is then scanned from left to right and every time a new subsequence of characters are encountered, a complexity counter is incremented by one. Predictable signals tend to have few distinct patterns resulting in lower number for LZC, while random signals exhibit higher values for LZC due to greater number of such distinct symbolic patterns.

LZC was computed for each EEG channel using the AntroPy toolbox [54], which implements median-based binarization prior to complexity estimation. Because raw LZC values are strongly influenced by sequence length, we employed a normalized variant of LZC [53], defined as

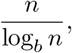

where *n* denotes the sequence length and *b* is the number of symbols in the discretized sequence *P*.

#### Detrended fluctuation analysis

Detrended fluctuation analysis captures long range temporal correlations in a time series and has been widely applied to human EEG data [55]. Briefly, detrended fluctuation analysis estimates the exponent/slope *α* describing the relation between fluctuations of the amplitude envelope obtained by Hilbert Transform and the temporal window size. The DFA scaling exponent *α* is the generalization of the Hurst exponent. Similar to Hurst exponent values, less than 0.5 indicate a white-noise process, while values larger than 0.5 suggest a correlated signal. However, DFA can return slopes larger than one in the case of unbounded, non-stationary signals like EEG [56] and are interpreted in terms of relative changes across conditions. We used the Antropy toolbox to compute the DFA scaling exponent [54].

#### Sample entropy

Sample entropy (SampEn) is a widely used measure of complexity of physiological time-series signals [57, 58]. Conceptually, SampEn quantifies the negative logarithm of the conditional probability that two sequences that are similar for *m* consecutive samples remain similar when one additional sample is included. Thus, lower SampEn values reflect greater predictability, whereas higher values indicate increased randomness or unpredictability of the signal.

Formally, a signal of length *N* is embedded into vectors of length *m*, yielding *N − m* + 1 template vectors. Pairwise similarity between template vectors is assessed using the Euclidean distance, and two vectors are considered similar when their distance is smaller than a tolerance threshold *r*. The SampEn value is then computed from the ratio of the number of similar vector pairs of length *m* + 1 to those of length *m*.

Sample entropy was computed using the NeuroKit2 toolbox [59] with an embedding dimension of *m* = 2, time delay *τ* = 1, and tolerance *r* = 0.2 times the standard deviation of the signal, corresponding to commonly used default parameters.

#### Spectral entropy

Spectral entropy (SpecEn) is a measure of the distribution of spectral power across frequencies and reflects the irregularity of the power spectrum [60]. It is computed as the Shannon entropy of the power spectral density. Higher SpecEn values indicate a more uniform distribution of spectral power across frequencies, whereas lower values reflect the concentration of power within specific frequency bands.

### Mental health assessment

Self-report data were acquired from campers: (1) in the week prior to or on the first day of camp; and (2) on the last day of camp. The self-report questionnaire asked campers about their demographics, sleep, ultra-processed food consumption, belonging, friendship, technology-habits, attitudes on being tech free (end of camp only) and the 47 MHQ items that span emotional, cognitive, and social functioning [61–63]. The 47 MHQ items, adapted from the adult MHQ [61–63] to ensure age-appropriate language and reading level have been similarly validated and are currently employed in the Youth Global Mind Study as well as in schools. In this study we considered only the MHQ items, along with the overall MHQ scores and the dimensional scores. The pre- and post-camp questionnaires can be found in the supporting information (S1 File and S2 File).

### Pre-post analysis

The individual participant was the unit of analysis, with pre- and post-camp measurements treated as paired observations within each participant.

For each subject, EEG features described above were computed separately for pre-camp (first day of camp) and post-camp (last day of camp) recordings and averaged across a fixed, subject-specific subset of epochs to ensure comparability across conditions. Furthermore, we averaged the EEG features across channels within frontal (AF3, F7, F3, F4, F8, AF4), central (C3, FC5, C4, FC6), parietal (P7, P8), and occipital (O1, O2) electrode groups.

For each subject, electrode group, and feature, EEG change scores were computed as

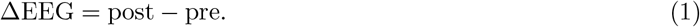

Change in various elements of mind health as well as aggregate scores were derived from the questionnaire items administered before and after the camp. Mind health change scores were computed analogously as

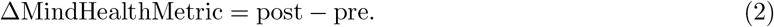

A mind health metric was considered to have changed for a subject if ΔMindHealthMetric ≠ 0, allowing for changes in either direction.

### Correspondence between EEG and mind health metrics

These EEG–mind health correspondence analyses were exploratory. To assess whether EEG changes beyond a meaningful magnitude were associated with positive changes in mind health metrics, we performed a threshold-based overlap analysis. For each EEG feature and region, EEG change scores were standardized (z-scored) across subjects to enable comparability across features. An EEG feature was considered to have changed if

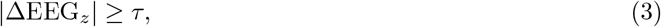

with *τ* = 0.5 standard deviations.

For each combination of mind health metric, EEG feature, and brain region, subjects were classified into a 2 *×* 2 contingency table based on whether the EEG change exceeded the threshold and whether the change in mind health metric was non-zero. From this table, three complementary overlap metrics were computed: precision, recall, and the Jaccard index. Precision quantified the probability of change in mind health metric given EEG change, recall quantified the probability of EEG change given a change in mind health metric, and the Jaccard index quantified the proportion of subjects showing both EEG and mind health change relative to all subjects showing either change. Analyses were performed separately for frontal, central, parietal, and occipital regions.

### Statistical analysis

Differences in EEG features before and after the camp were assessed using the Wilcoxon signed-rank test. Effect sizes were quantified using Cliff’s delta (Cliff’s *δ*), defined as

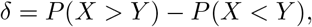

where *X* and *Y* denote observations from the two paired conditions. Statistical tests were two-sided, with *p <* 0.05 considered statistically significant. Given the exploratory nature of the EEG analyses, no correction for multiple comparisons was applied; individual feature–region comparisons should therefore be interpreted as exploratory. No imputation of missing data was performed. Analyses involving mind health measures were restricted to participants with the corresponding pre- and post-camp data available. All statistical analyses were performed using Python version 3.13

## Results

The final EEG sample consisted of 30 adolescents (23 from the 2024 cohort and 7 from the 2025 cohort). Participant demographic and baseline characteristics are summarized in Table 1.

**Table 1.** Baseline characteristics of the EEG study sample.

| Characteristic | EEG sample ($N = 30$) |
| --- | --- |
| Age, years, mean ± SD<sup>a</sup> | 14.6 ± 1.1 |
| Biological sex<sup>b</sup> |  |
| Male, $n$ (%) | 14 (53.8%) |
| Female, $n$ (%) | 12 (46.2%) |

<sup>a</sup> Exact numerical age was available for 25 participants; one additional participant was recorded as "Younger than 13," and age was unavailable for four participants. Percentages for biological sex are calculated among participants with available data ( $n = 26$ ).
<sup>b</sup> Biological sex was available for 26 of the 30 participants.

We compared the changes in features computed from EEG signals recorded before and after the camp. These included DFA scaling exponent, aperiodic slope and offset, LZC, SampEn, SpecEn and spectral power in *δ*, *θ*, *α* and *β* bands. Fig 3 illustrates the dominant direction of change in these EEG features between the beginning and end of the camp, averaged over electrode groups — frontal (AF3, F7, F3, F4, F8, AF4), central (C3, FC5, C4, FC6), parietal (P7, P8) and occipital (O1, O2). Overall multiple metrics changed significantly, particularly in the occipital and central regions.

**Fig 3.**
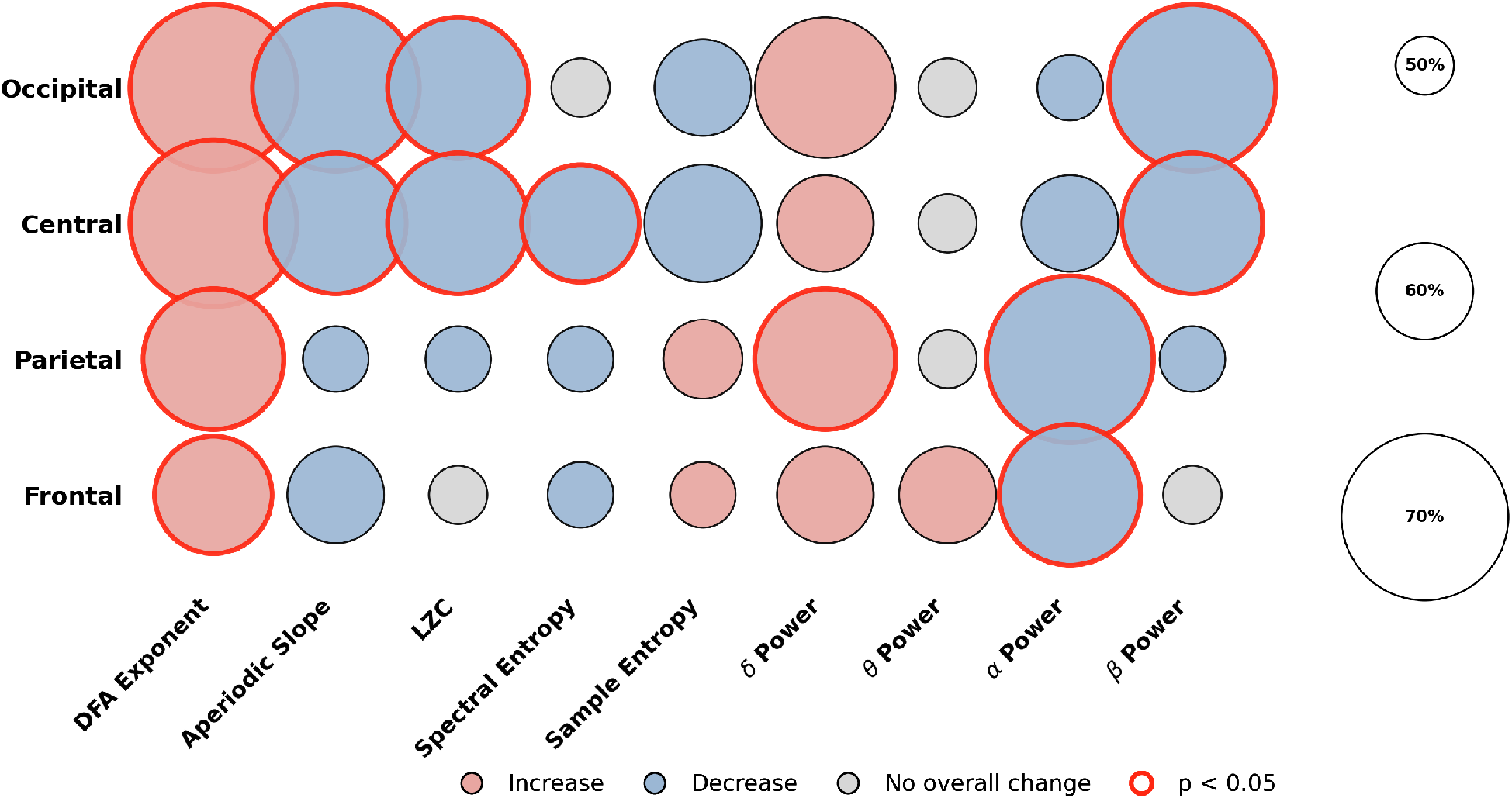
Dominant direction of change in EEG features after camp. Bubble plots summarize within-subject pre–post changes across scalp regions (rows; Frontal, Parietal, Central and Occipital) and EEG features (columns; DFA exponent, aperiodic slope, LZC, spectral entropy, sample entropy, and band power in *δ*, *θ*, *α*, and *β*). Bubble area is proportional to the percentage of participants showing an increase (light red) or decrease (light blue) following camp. Gray bubbles indicate an even split (50% increase, 50% decrease). Red outlines denote feature–region pairs with a statistically significant pre–post difference (Wilcoxon signed-rank test, p *<* 0.05).

### Changes in EEG complexity

Overall, following camp participation there was a shift towards more structure or less randomness in the EEG signal, which was most prominent and significant in the channels across occipital and central regions.

The DFA scaling exponent increased robustly across all channel groups, with 63–70% of subjects showing an increase depending on the region (Wilcoxon signed-rank test, p *<* 0.01 for all regions). Other metrics, where the direction of increased structure or decreased randomness corresponds to a lower value, opposite that of the DFA exponent, were decreased post camp. The aperiodic slope decreased 60–70% with statistically significant effects restricted to occipital and central channels (p *<* 0.05). LZC and spectral entropy showed consistent post-camp decreases, with 57–67% and 53–63% of subjects showing reductions respectively, and statistically significant effects in central and occipital channels for LZC (p *<* 0.05) and central channels for spectral entropy (p *<* 0.05). Sample entropy also decreased in central and occipital regions in 53–63% of subjects, but these changes did not reach significance; in frontal and parietal regions, 53–57% of subjects showed a post-camp increase, also non-significant. Fig 4 shows subject-wise distributions for measures reaching statistical significance (p *<* 0.05; see also Fig 3), averaged across electrode groups.

**Fig 4.**
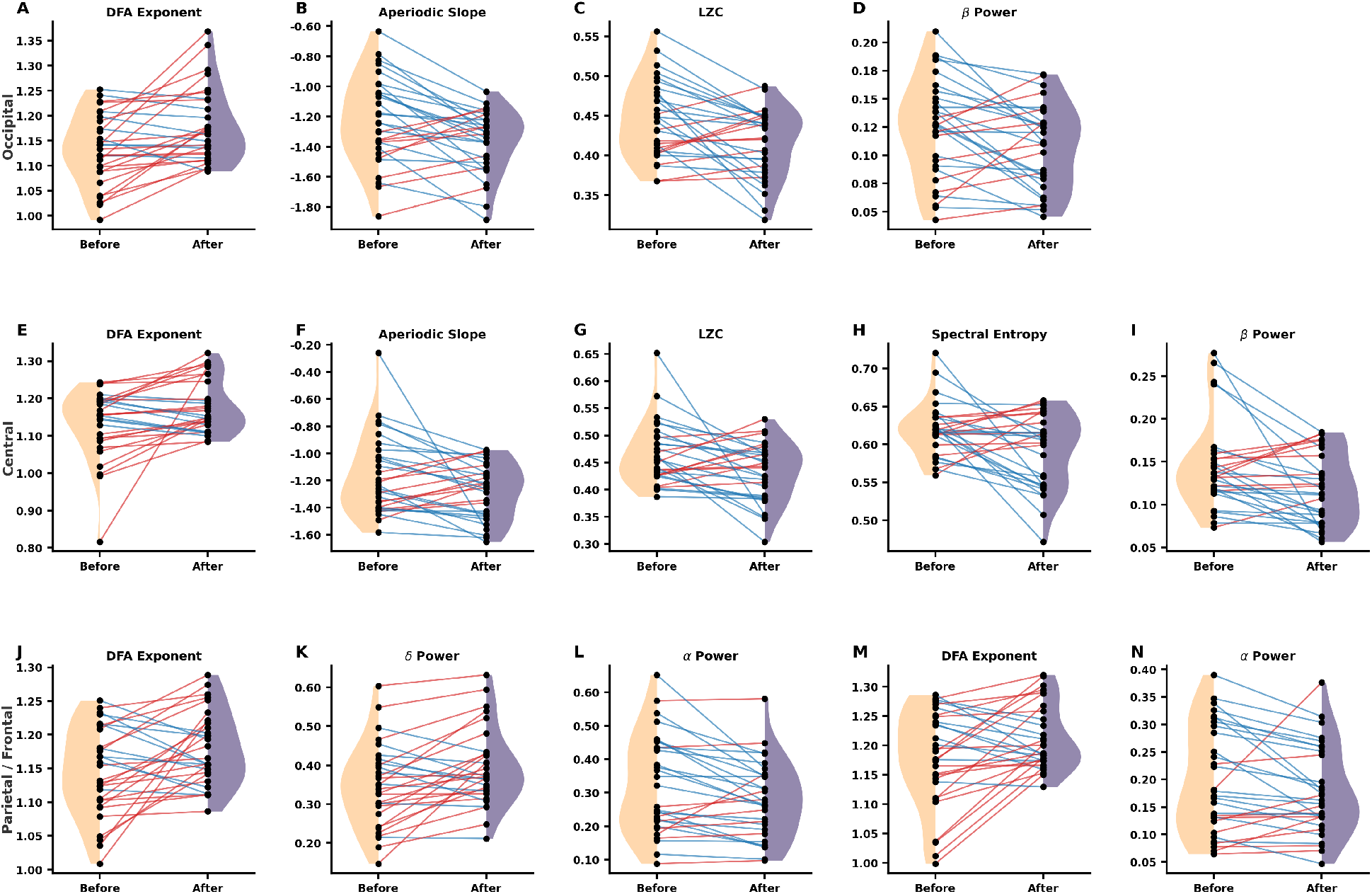
Subject-wise pre–post changes in EEG features for significant region–feature pairs (Wilcoxon signed-rank test, p *<* 0.05). Panels show paired measurements before and after camp for each participant (black points), connected by lines (red, increase; blue, decrease). Half-violin kernel density estimates summarize the marginal distributions at each time point (Before, left; After, right). (A–D) Occipital—DFA exponent, aperiodic slope, LZC, and *β* power; (E–I) Central—DFA exponent, aperiodic slope, LZC, spectral entropy, and *β* power; (J–L) Parietal—DFA exponent, *δ* power, and *α* power; (M and N) Frontal—DFA exponent and *α* power.

A complete summary of the complexity measures, including corresponding p-values and Cliff’s *δ* effect sizes, is provided in the supporting information (S3 File).

### Changes in EEG spectral power

Overall, EEG spectral power showed a shift toward lower frequencies following the camp, consistent with steeper aperiodic slopes and increase in signal predictability or structure. Among the spectral power measures computed in four key frequency bands, power in the *δ* and *θ* bands increased post-camp, whereas *α* power decreased across all electrode groups. In contrast, *β* power exhibited post-camp decreases restricted to the occipital and central channels, while in the frontal and parietal channels, equal numbers of subjects exhibited increases and decreases in *β* power. We observed a statistically significant post-camp increase in *δ*-band power in parietal channels (Wilcoxon signed-rank test, p *<* 0.05), with 70% of subjects showing higher values after the camp whereas *α* power decreased significantly in the frontal and parietal channels (67% and 70% of subjects, respectively), and *β* power decreased significantly in the occipital and central regions (67% and 63% of subjects, respectively). Fig 4 shows the subject-wise distributions of EEG spectral measures that exhibited statistically significant pre–post differences (Wilcoxon signed-rank test, p *<* 0.05), averaged across electrode groups. A complete summary of the spectral results, including corresponding values and Cliff’s *δ* effect sizes, is provided in the supporting information (S3 File).

### Changes in mind health metrics

Camp participation was associated with broad improvements in self-reported mind health assessed through various capacities, problems, and dimensional scores that aggregated across subsets of capacities and problems (see S3 File). Given that pre and post EEG recordings and mind health metrics were available for 23 of the 130 camp participants, statistical power was limited, and conventional significance thresholds were not appropriate as the primary selection criterion. We therefore adopted a two-stage criterion for reporting: measures were included if at least 40% of participants showed change in the expected direction and the Wilcoxon signed-rank test yielded p *<* 0.4. This approach captures meaningful trends that would be obscured by strict p-value thresholds in a small sample. The majority of selected measures using this criteria also reached conventional significance (*p <* 0.05) in the larger sample of 192 participants, validating that the observed improvements reflect reliable camp effects.

Four capacity measures met this criterion: emotional resilience, focus and concentration, planning and organization, and sleep quality. Of these, emotional resilience showed the largest and most consistent improvement, reaching conventional statistical significance (p *<* 0.05), with 43% of participants showing improvement. Focus and concentration and sleep quality each showed improvement in 52% of participants (p = 0.14 and p = 0.36, respectively), while planning and organization improved in 43% of participants (p = 0.22).

Among problem measures, restlessness and hyperactivity showed the most pronounced change, with 52% of participants showing reduction after camp and a statistically significant Wilcoxon test (p *<* 0.05). Three additional problem measures — confusion and slowed thinking (43%, p = 0.27), fear and anxiety (48%, p = 0.31), and detachment from reality (43%, p = 0.39) — showed trends in the expected direction consistent with reduced scores following the intervention. It is to be noted that planning and organization and confusion and slowed thinking did not reach significance in the larger sample of 130 participants but were retained as both met the selection criteria in the EEG subsample and contribute to a more complete characterization of the mind health metrics accompanying the EEG shifts reported below.

Composite dimension scores showed the most consistent pattern of improvement. Mind Body Connection and Adaptability and resilience each improved in 70% of participants (p = 0.05 and p = 0.19 respectively), while the overall MHQ score improved in 65% of participants (p = 0.26). Cognition and Mood and outlook each improved in 61% of participants (p = 0.31 and p = 0.32). Although these dimension-level changes did not reach conventional significance, the high proportion of participants showing improvement — consistently above 60% — suggests a broad and directionally coherent shift in mind health following camp participation.

Taken together, these findings indicate that participation in the device-free camp was associated with improvements spanning multiple domains of mind health, with the most statistically reliable effects observed for emotional resilience and restlessness.

### Covariance of changes in EEG features and mind health metrics

To examine whether the changes in EEG and mind health metrics observed after camp were related, we assessed the co-variation between pre-post changes in each EEG feature and each mind health metric. For each subject and each EEG feature–mind health metric pair, we determined whether the two measures changed in a concordant direction, i.e., whether the EEG shifted toward a more structured or low-frequency dominant state at the same time as the mind health metric improved.

Figs 5 and 6 display the percentage of subjects showing this concordant pattern for each combination of EEG feature, mind health metric, and scalp region.

**Fig 5.**
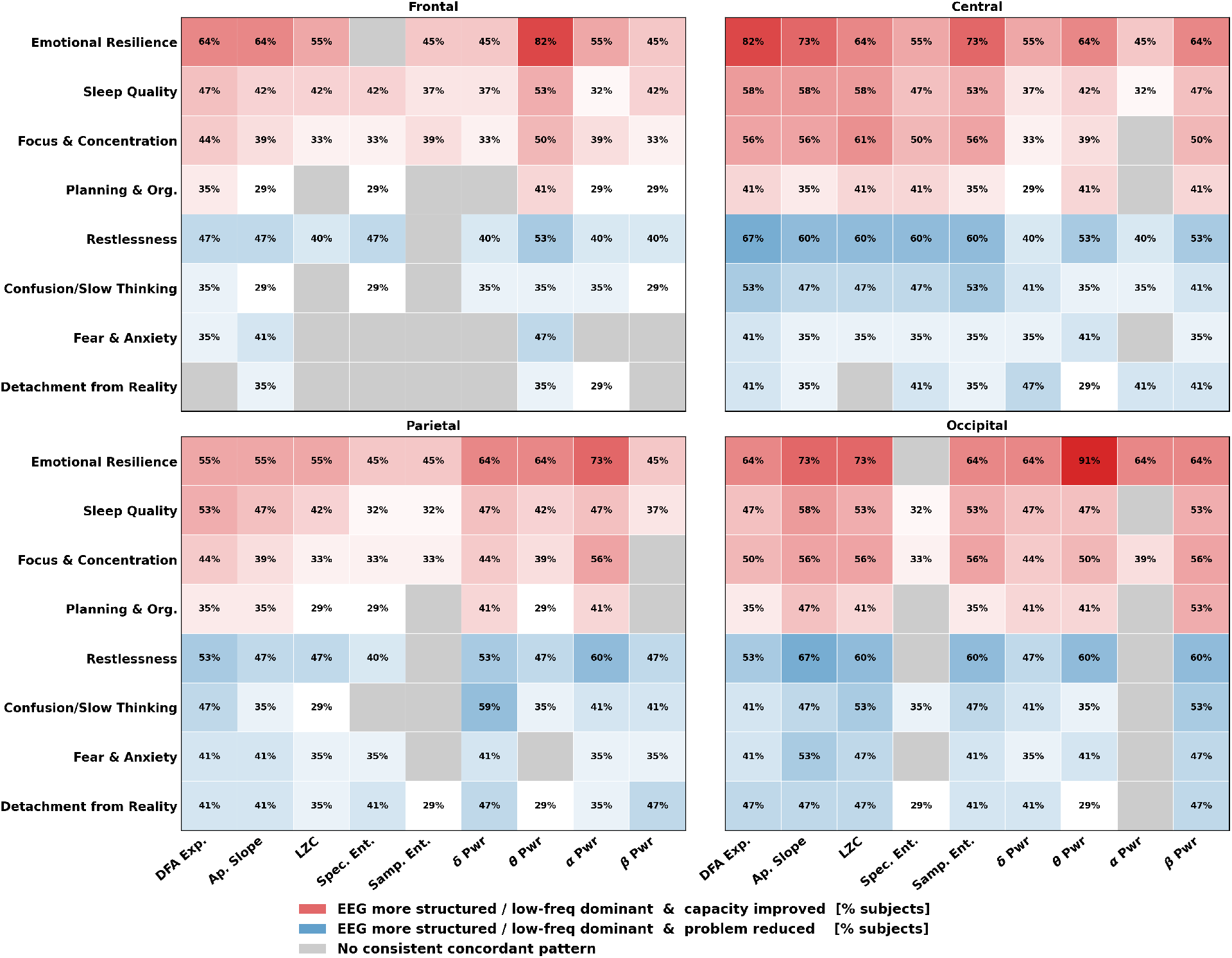
Co-variation between pre-post changes in EEG features and capacity and problem measures across four scalp regions. Each cell displays the percentage of subjects showing a concordant pattern between EEG and reported change in mind health. Red cells indicate that the EEG shifted toward a more structured or low-frequency dominant state, meaning the DFA exponent, *δ* power, or *θ* power increased, or the aperiodic slope steepened, or LZC, spectral entropy, sample entropy, *α* power, or *β* power decreased, concurrent with improvement in a capacity measure (higher score). Blue cells indicate the same EEG shift concurrent with reduction in a problem measure (lower score reflecting improvement). Gray cells indicate no consistent concordant pattern. Color saturation scales with the percentage of subjects showing the concordant pattern. EEG features shown on the x-axis are DFA exponent (DFA Exp.), aperiodic slope (Ap. Slope), Lempel–Ziv complexity (LZC), spectral entropy (Spec. Ent.), sample entropy (Samp. Ent.), *δ*, *θ*, *α*, and *β* band power. Since four directional combinations are possible, the chance-level expectation for any single quadrant is 25%.

**Fig 6.**
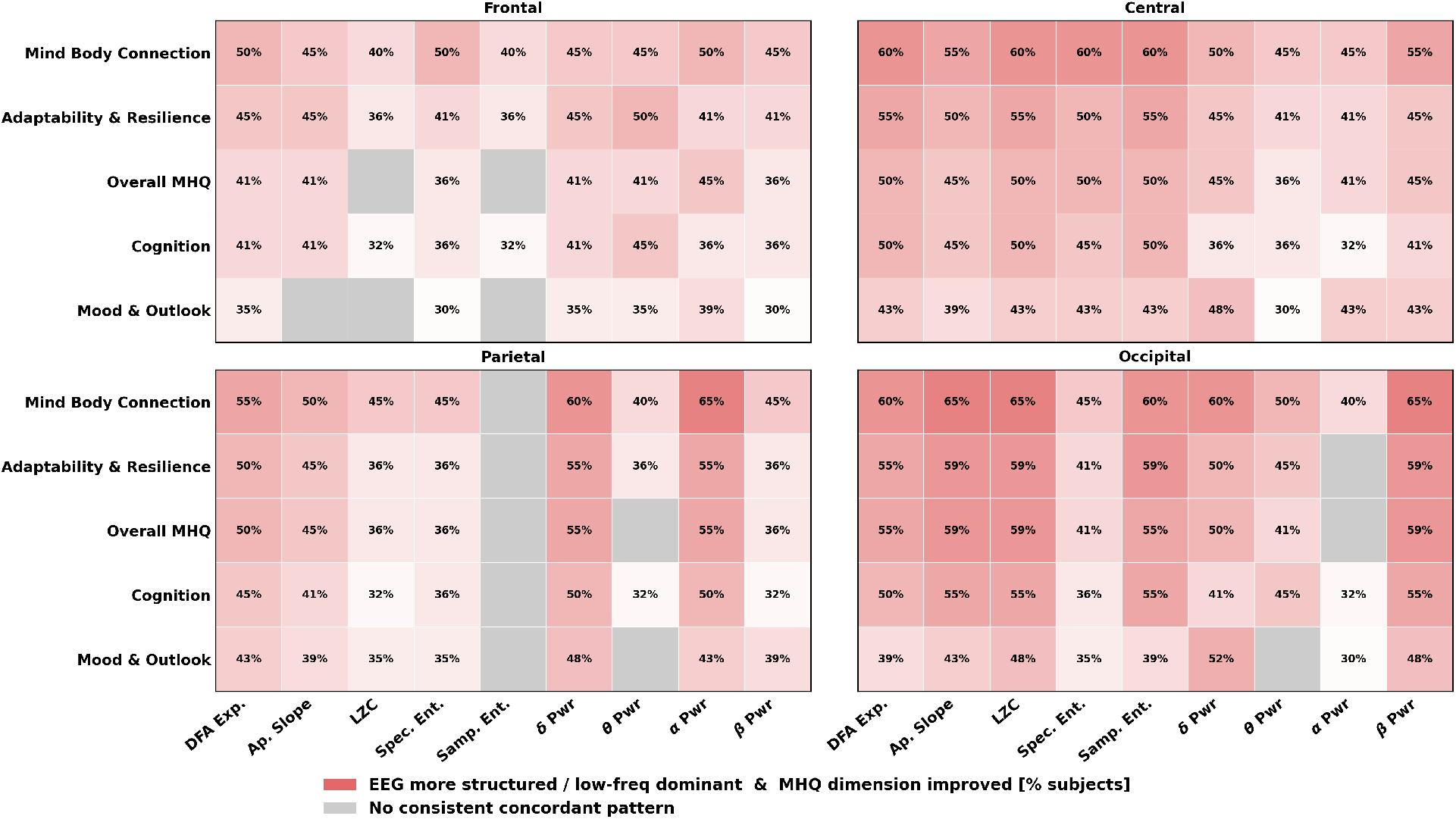
Co-variation between pre-post changes in EEG features and composite MHQ dimension scores across four scalp regions. Each cell displays the percentage of subjects showing a concordant pattern between EEG and reported change in mind health. Red cells indicate that the EEG shifted toward a more structured or low-frequency dominant state concurrent with improvement in a composite dimension score. Gray cells indicate no consistent concordant pattern. Color saturation scales with the percentage of subjects showing the concordant pattern. Dimension measures shown on the y-axis are Mind-Body Connection (MBC), Adaptability and resilience, overall Mental Health Quotient (MHQ), Cognition, and Mood and outlook. EEG features on the x-axis are as in Fig 5. Since four directional combinations are possible, the chance-level expectation for any single quadrant is 25%.

Since four directional combinations are possible — (1) EEG shifts towards more structured state and mind health metrics improve, (2) EEG shifts towards more structured state and mind health metrics worsen, (3) EEG shifts towards more random state and mind health metrics improve, and (4) EEG shifts towards more structured state and mind health metrics worsen — random expectation for any single quadrant is 25%.

Red cells indicate that the EEG shifted toward a more structured or low-frequency dominant state, meaning the DFA exponent, *δ* power, or *θ* power increased, or the aperiodic slope, LZC, spectral entropy, sample entropy, *α* power, or *β* power decreased, concurrent with increase in a capacity or dimension measure. Blue cells indicate the same EEG shift concurrent with decrease in a problem measure. Gray cells indicate no consistent concordant pattern. Deeper color saturation indicates a higher percentage of subjects showing the concordant pattern. The concordant percentages reported throughout consistently exceed chance, and the remaining subjects are distributed across three non-concordant patterns rather than uniformly opposing the dominant direction.

Among capacity measures, emotional resilience showed the strongest and most spatially widespread concordance with EEG change (Fig 5). In the occipital region, 91% of subjects showed a concurrent increase in *θ* power alongside improvement in emotional resilience. In central channels, the DFA exponent increase co-occurred with emotional resilience improvement in 82% of subjects, while decrease in the aperiodic slope and reduction in LZC co-varied with the same mind health metric in 73% and 64% of subjects respectively. Focus and concentration showed consistent concordance with DFA exponent, aperiodic slope, and LZC changes in 50–61% of subjects, predominantly in central and occipital channels. Sleep quality and planning and organisation showed more moderate concordance, generally in the 35–58% range across regions and features.

Among problem measures, restlessness showed the most consistent concordant pattern (Fig 5). In central channels, 60–67% of subjects simultaneously showed EEG shifts toward more structured dynamics and reduced activation — including increases in DFA exponent, steepening of the aperiodic slope, and reductions in LZC, spectral entropy, and sample entropy — and reductions in restlessness. Occipital channels showed a similar pattern, with aperiodic slope steepening and reductions in LZC and *β* power co-varying with restlessness reduction in 60–67% of subjects. Confusion and slowed thinking showed notable concordance particularly in parietal channels, where 59% of subjects showed co-occurring increases in *δ* power and reported mind health improvement. Fear and anxiety and detachment from reality showed weaker and less spatially consistent concordance, with most cells in the 35–47% range.

The composite dimension measures showed a broadly consistent red pattern, indicating that EEG shifts toward more predictable dynamics co-occur with improvements in Mind-Body Connection, Adaptability and resilience, overall MHQ, Cognition, and Mood and outlook in a substantial proportion of subjects (Fig 6). Mind-Body Connection showed the strongest concordance overall, with 60–65% of subjects in central and occipital regions showing co-occurring EEG structural shifts, including increases in DFA exponent and reductions in LZC and *β* power, alongside improvement in this composite score. Adaptability and resilience and overall MHQ showed similar patterns, with concordance of 55–59% in occipital channels across multiple EEG features. Cognition and Mood and outlook showed slightly weaker but directionally consistent concordance. Notably, gray cells, indicating the absence of a consistent concordant pattern, were rare across the dimensions panel, suggesting that the relationship between EEG structural change and mind health improvement was broadly consistent regardless of which specific EEG feature was examined.

Overall, central and occipital regions consistently showed the strongest and most frequent concordant patterns, while frontal concordance was generally lower. This regional trend is consistent with the group-level EEG results reported above (see Fig 3), and suggests that the neural correlates of self-reported mind health metrics following camp participation are most reliably observed in posterior and central scalp regions. The interpretation of regional specificity is however constrained by the limited spatial resolution of scalp EEG, and source-level analyses would be needed to identify the specific cortical generators underlying these patterns.

### Metrics of EEG-mind health concordance

As this is a pilot study with a small N, we quantified the correspondence between EEG features and mind health metrics in each group of scalp electrodes, using precision, recall, and Jaccard indices (see Figs 7 and 8 for precision and S3 File for recall and Jaccard).

**Fig 7.**
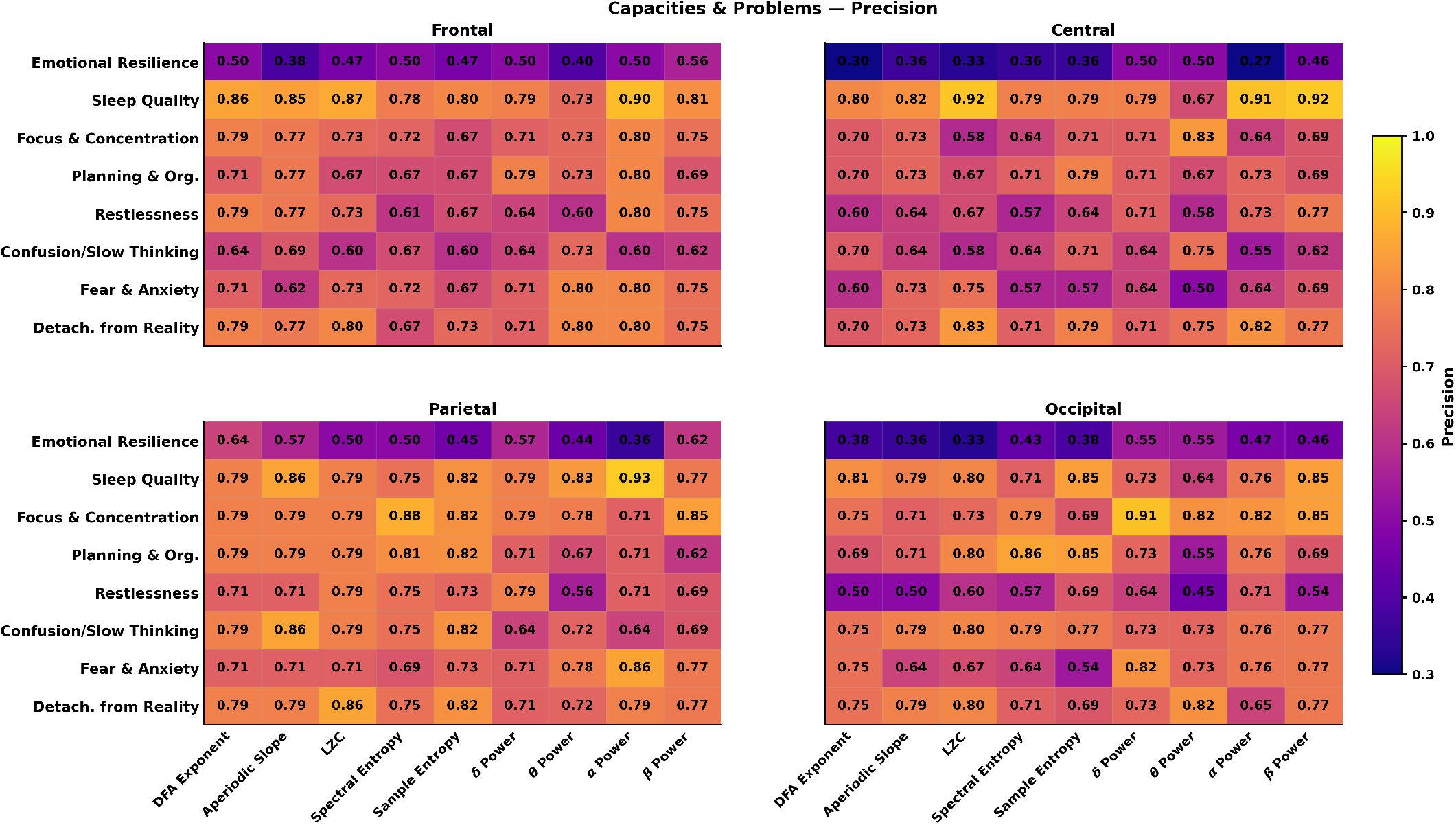
Precision of EEG-mind health change across brain regions for mind health metrics across capacities and problems. Precision values reflect the proportion of participants showing EEG changes beyond a defined threshold who also exhibited corresponding change in the reported mind health metrics. Each panel corresponds to a scalp region (frontal, central, parietal, occipital), with EEG features on the x-axis and mind health metrics on the y-axis, and precision values indicated by color.

**Fig 8.**
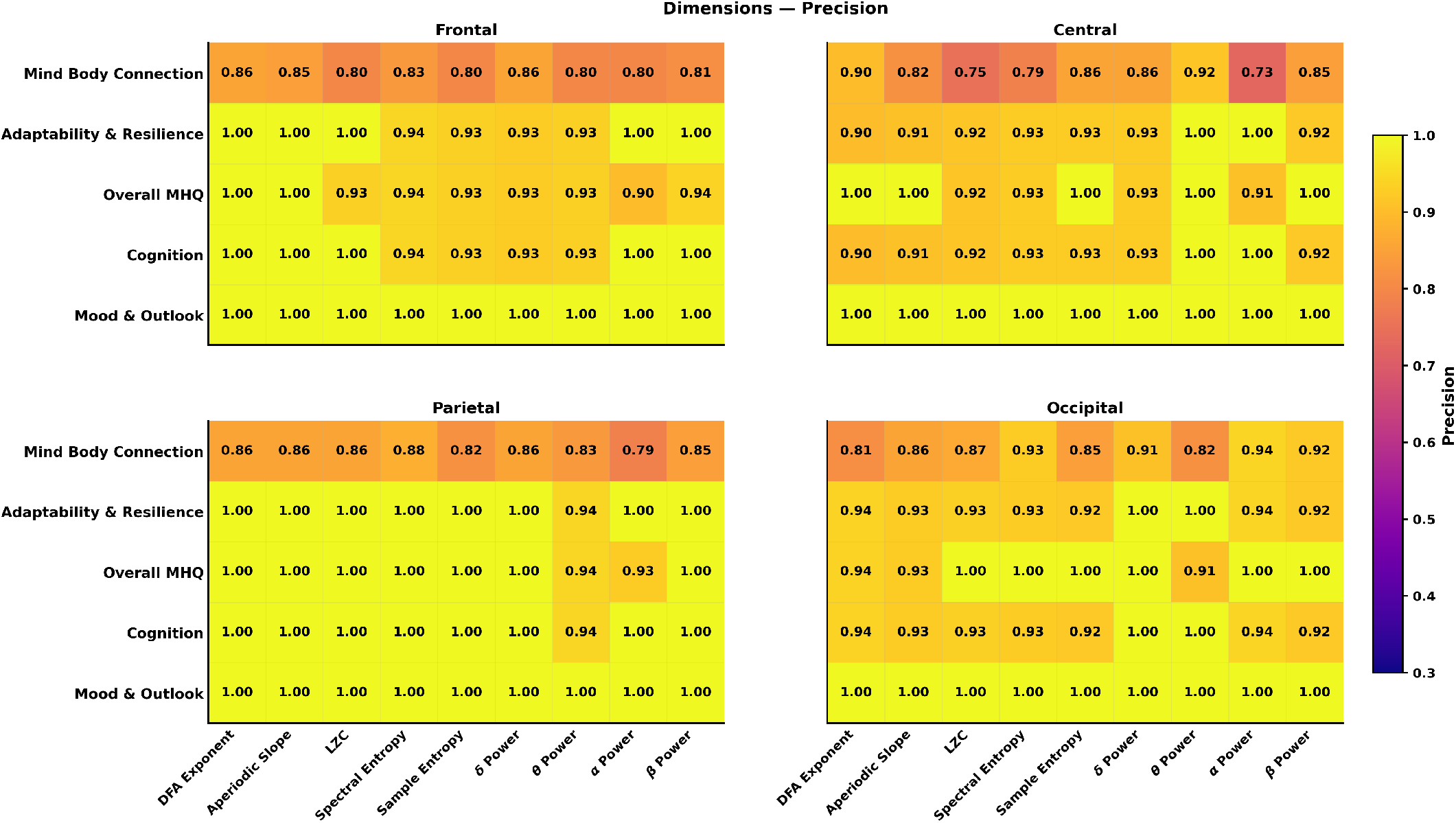
Precision of EEG-mind health change across brain regions for mind health metrics across dimensions. Precision values reflect the proportion of participants showing EEG changes beyond a defined threshold who also reported corresponding change in mind health metric. Each panel corresponds to a scalp region (frontal, central, parietal, occipital), with EEG features on the x-axis and mind health metrics on the y-axis, and precision values indicated by color.

Overall, overlap patterns were spatially structured, with parietal and occipital regions generally exhibiting higher EEG–mind health concordance than frontal and central regions.

Precision values were consistently high across EEG–mind health metric pairs, ranging between 0.9–1.0 for EEG–dimension pairs (meaning that 90–100% of those who had a significant change in the EEG reported a significant change in the mind health metric) and 0.7–0.9 for EEG–capacity and EEG–problem pairs. The exception was emotional resilience, where precision was comparatively lower (0.4–0.6), suggesting that not all subjects who showed EEG changes also reported improvements in emotional resilience. In parietal and occipital regions, precision frequently exceeded 0.8 and in several cases approached 1.0, indicating that subjects exhibiting EEG changes beyond the threshold were highly likely to show corresponding improvements in perceived mind health. Precision values were comparatively lower in frontal and central regions, though still consistently above chance, suggesting that EEG changes in these regions were less specific to concurrent changes in perceived mind health.

Recall values revealed a complementary pattern. While frontal and central regions showed moderate recall, parietal and occipital regions consistently demonstrated higher recall (0.7–0.82), indicating that perceived improvements in mind health were more often accompanied by detectable EEG changes in posterior regions. Notably, emotional resilience showed a pattern of low precision but relatively high recall, suggesting that while EEG changes were not reliably specific to emotional resilience improvements, the majority of subjects who did report emotional resilience improvements showed concurrent EEG changes. This suggests that improvements in emotional resilience were consistently accompanied by EEG changes, but the EEG changes themselves were not specific to emotional resilience, pointing to a concurrent but non-specific effect that the EEG measures examined here do not fully capture.

The Jaccard index integrated both precision and recall, providing a conservative measure of EEG–mind health overlap. Jaccard values were highest in frontal, parietal and occipital regions (0.6–0.8), especially for measures of signal predictability, including the DFA scaling exponent, aperiodic slope, and spectral entropy, as well as for *α*- and *θ*-band power. Across multiple mind health metrics, these posterior/frontal EEG features exhibited Jaccard values exceeding 0.6, indicating substantial overlap between subjects showing EEG changes and improvements in mind health.

Together, these results demonstrate that EEG changes exceeding a modest, standardized threshold reliably co-occur with a perceived change in mind health, with the strongest correspondence observed in posterior brain regions.

## Discussion

This study provides the first characterization of how changes in neural dynamics associated with participation in a device-free residential summer camp in adolescents correspond with perceived changes in select mental capacities and problems as well as aggregate mind health and dimensional composites. We found that multiple EEG measures changed significantly following the intervention, and that these changes co-varied with improvements in mind health metrics. Taken together, the findings demonstrate a shift toward greater signal predictability with high precision, particularly in the occipital and parietal regions, across complexity measures, and a shift toward lower-frequency spectral activity. Both patterns are consistent with a less excitable or more structured baseline neural state following three weeks of reduced digital stimulation and sustained nature exposure. Notably, reductions in restlessness and hyperactivity and improvements in focus co-varied most consistently with these neural changes — outcomes that are themselves well-documented consequences of nature exposure [20], suggesting the observed neural shifts may reflect the same restorative processes proposed by Attention Restoration Theory and Stress Recovery Theory.

### Spectral changes are consistent with calmer brain state

EEG studies have shown that exposure to nature reduces *β* activity, which is associated with lower stress and cognitive load [35, 64, 65]. In line with these findings, we observed a significant reduction in *β* power following the camp intervention. At the subject level, this reduction co-varied most consistently with improvements in restlessness, emotional resilience, and sleep quality, as well as with the composite dimension measures of Mind-Body Connection, Adaptability and resilience, and overall MHQ, predominantly in central and occipital channels. The reduction in *β* power across all scalp regions was also consistently associated with reductions in restlessness, suggesting that the reduction in *β* power may reflect the neural basis of the observed improvements in attentional regulation and restlessness following the intervention. Precision analyses further confirmed the specificity of this relationship. Subjects who showed *β* power reduction beyond a defined threshold also showed corresponding improvements in restlessness and mind health metrics across dimensions (see Figs 7 and 8).

The *α* power during eyes-closed rest was also reduced after camp. While *α* oscillations are typically prominent during relaxed wakefulness, their magnitude is known to vary substantially with environmental context and lifestyle factors. Prior work has demonstrated that chronic exposure to modern technological environments elevates resting *α* power, suggesting it may reflect a baseline state of heightened neural stimulation rather than pure relaxation [66]. Under this interpretation, the reduction in *α* power observed here may reflect a recalibration of resting neural baseline following sustained removal from digital stimulation, rather than a paradoxical increase in arousal. Consistent with this, *α* reduction co-varied with improvements in emotional resilience, focus and concentration, and the composite dimension measures in most scalp regions, though this association was less consistent in occipital regions compared to *β* power. Reductions in *α* power in parietal channels were also associated with improvements in focus and concentration in 56% of subjects, suggesting a link between reduced high-frequency neural idling and improved attentional capacity following the intervention.

The reduction in *α* and *β* power was correspondingly accompanied by an increase in *δ* and *θ* activity. In eyes-closed resting EEG, increased slow-frequency activity is commonly associated with reduced vigilance and lower levels of cortical activation. The increase in *θ* power showed particularly strong concordance with improvements in emotional resilience, most prominently in occipital channels. It is worth noting that concordance percentages reflect the proportion of subjects with non-zero change on both the EEG and reported mind health metric rather than the full sample; consequently, high concordance values can emerge even when the group-level EEG change is modest. In this case, the high concordance between *θ* power and emotional resilience reflects strong directional agreement among subjects who showed any change on both measures, rather than implying a large-scale *θ* increase across all participants. Increases in *δ* power were associated with improvements in Adaptability and resilience, Cognition, and Mind-Body Connection in 40% or more subjects, reflecting the broader shift toward slow-frequency dominance following the intervention.

Together, these spectral changes including reductions in *α* and *β* power alongside increases in *δ* and *θ* power were reflected in steeper aperiodic slopes, increased DFA scaling exponents, and reduced Lempel–Ziv complexity. Collectively, these findings suggest that the camp intervention was associated with a shift toward slower, more predictable neural dynamics and reduced cortical activation, which may reflect a calmer baseline brain state following a period of reduced digital stimulation and increased exposure to natural environments.

### EEG complexity measure changes reflect reduced excitation

Following camp participation, DFA scaling exponents increased robustly across all scalp regions, while LZC, spectral entropy, sample entropy, and the aperiodic slope decreased, particularly in central and occipital regions. Although these measures change in opposite numerical directions, they reflect the same underlying shift: the neural signal became more structured and less random. The DFA exponent indexes long-range temporal correlations where higher values indicate that neural activity is more organised across time, while lower entropy, LZC, and sample entropy values indicate a more predictable signal. A steeper aperiodic slope similarly reflects a greater relative dominance of lower frequencies, indicative of signal predictability.

These neural changes co-varied with improvements across multiple mind health factors. Among capacity measures, improvements in emotional resilience, focus and concentration, sleep quality, and planning and organisation were accompanied by shifts toward more predictable EEG dynamics, predominantly in central and occipital channels. Among problem measures, reductions in restlessness and hyperactivity showed the most consistent co-variation with shifts in EEG complexity measures in direction of more predictable signals, followed by reductions in confusion and slowed thinking, fear and anxiety, and detachment from reality. Mind-Body Connection, Adaptability and resilience, overall MHQ, Cognition, and Mood and outlook also showed broadly consistent co-variation with EEG complexity changes across all scalp regions.

Within the excitation–inhibition (E/I) balance framework, steeper aperiodic slopes and increased DFA scaling exponents have been associated with relatively greater inhibitory influence or reduced cortical excitation [67–69]. This interpretation is further supported by the pattern observed for LZC. Prior research suggests that lower LZC is associated with a predominance of inhibitory over excitatory neural activity [70, 71], and a recent study found that caffeine — which promotes cortical activation by blocking adenosine receptors — increased LZC during sleep [56], consistent with the opposite pattern observed here. Collectively, these findings suggest that the camp intervention shifted baseline brain dynamics toward a less excitable, more structured state, which may underlie the observed improvements across the range of mind health metrics reported here.

### Neural and mind health metrics are linked

While the absence of a control group means we cannot establish causality, several features of the data strengthen a causal interpretation. The precision analyses demonstrated that EEG changes were not merely widespread but were specifically co-localised with changes in mind health metrics. Participants who showed EEG shifts were substantially more likely to also show corresponding improvements across specific capacities and problems, and composite dimension measures, particularly in posterior and central regions. This individual-level linkage is consistent with population-level findings from a larger sample of the same camp, which similarly reported significant pre-post improvements in self-reported mind health following device-free camp participation [32]. It is also to be noted that the changes in primary capacities and problems are related — less restlessness and greater focus have been related to improved sleep quality.

However, the relationship was not symmetric. EEG changes were more reliably accompanied by improvements in mind health metrics than the reverse. Some participants may have reported mind health improvements without corresponding EEG shifts, which could reflect genuine individual variability in response to the intervention, or a tendency to rate items more positively due to a general halo effect associated with an enjoyable camp experience.

Disentangling these possibilities would require a control condition and more objective measures of mind health.

### Limitations

As a pilot study, the present findings establish the feasibility of detecting measurable, environmentally-linked EEG changes in a real-world adolescent sample, laying the groundwork for larger controlled studies to confirm and extend these results. A few limitations of our study are worth considering. First, the absence of a control group limits causal inference. Without a group of adolescents continuing their typical digital, indoor dominant lifestyle over the same period, it is not possible to rule out the influence of other factors on the observed changes such as a decreased stress level due to being out of school. However, we do note that the camp session was held in July when students had already been out of the school environment for a month or more. Second, the camp intervention is inherently multifaceted — it simultaneously removed digital devices, increased nature exposure, restructured social interaction, altered sleep schedules, and introduced physical activity. This makes it difficult to attribute any single component of the camp experience to the observed changes in neural and mind health metrics. Third, scalp EEG has limited spatial resolution, constraining interpretation of the regional patterns we observed here. Thus our results cannot be confidently attributed to any specific underlying neural generators, and source-level analyses or higher-density EEG would be needed to address this. Fourth, the sample was relatively small and drawn from a single residential camp. Families who choose device-free nature-rich camps may also differ systematically from the broader adolescent population in terms of attitudes or access toward technology, socioeconomic status, or baseline mind health, further limiting the generalizability of our findings. Nonetheless, our results point to concomitant shifts in neurophysiology and reported mind health associated with the camp environment.

## Conclusions

This study provides the first evidence that participation in a device-free, nature-rich residential summer camp is associated with coordinated changes in both neural dynamics and reported mind health in adolescents. More generally, it is evidence of the profound impact of even short term shifting of the environment on brain physiology and corresponding perception of mind health. Following three weeks at camp, participants exhibited a consistent pattern of neural change during a resting state — including steeper aperiodic slopes, decreased offsets, increased DFA scaling exponents, reduced Lempel-Ziv complexity and entropy, and decreased alpha and beta power — alongside improvements in restlessness, hyperactivity, focus, sleep, and broader measures of mind health. Taken together, these findings are consistent with a shift toward a calmer, more structured baseline neural state, characterized by relatively greater inhibitory influence and reduced cortical excitation. Critically, these neural changes were not independent of reported changes in mind health. Instead, they co-varied with the improvements in mind health metrics, particularly in posterior and central scalp regions. These findings extend prior behavioral work on device-free camp settings by demonstrating that the benefits of such environments are reflected not only in self-reported changes in mind health but in measurable shifts in underlying brain dynamics. More broadly, they suggest that sustained immersion in nature-rich, device-free environments may support adolescent neural and psychological health in ways that brief laboratory exposures or individual digital detox efforts cannot replicate. Future work incorporating control conditions, longitudinal follow-up, and higher spatial resolution methods will be needed to establish causality and identify the specific mechanisms driving these changes.

## Data Availability

All the codes related to the publication will be made available via github.

## Supporting information

**S1 Protocol. Study protocol.** Protocol for the study evaluating the impact of a device-free residential summer camp on adolescent mental health and wellbeing.

**S2 File. Pre-camp survey questionnaire (MHQ).** Full 47-item Mind Health Quotient questionnaire administered to campers prior to or on the first day of camp.

**S3 File. Post-camp survey questionnaire (MHQ).** Full 47-item Mind Health Quotient questionnaire administered to campers on the last day of camp.

**S4 File. Supplementary results.** Complete summary tables of complexity and spectral power EEG measures across scalp regions (pre–post camp), including p-values and Cliff’s *δ* effect sizes. Also includes bar plots of self-reported mind health metrics before and after camp, and heatmaps of recall and Jaccard index values for EEG–mind health metric overlap across brain regions.

## Acknowledgments

The authors thank the campers and families of Maine Teen Camp for their participation in this study, and the camp staff for their support during data collection.

## References

1. Twenge JM, Martin GN, Campbell WK. Decreases in Psychological Well-Being Among American Adolescents After 2012 and Links to Screen Time During the Rise of Smartphone Technology. Emotion. 2018;18(6):765–80. Available from: 10.1037/emo0000403. doi:10.1037/emo0000403.

2. Centers for Disease Control and Prevention. Youth Risk Behavior Survey Data Summary & Trends Report: 2009–2019. Centers for Disease Control and Prevention; 2020. Available from: https://www.cdc.gov/healthyyouth/data/yrbs/pdf/YRBSDataSummaryTrendsReport2019-508.pdf.

3. O’Reilly M, Dogra N, Whiteman N, Hughes J, Eruyar S, Reilly P. Is social media bad for mental health and wellbeing? Exploring the perspectives of adolescents. Clinical Child Psychology and Psychiatry. 2018 May;23(4):601–613. Available from: 10.1177/1359104518775154. doi:10.1177/1359104518775154.

4. Office of the Surgeon General. Social Media and Youth Mental Health. Washington, DC: U.S. Department of Health and Human Services; 2023. U.S. Surgeon General’s Advisory. Available from: https://www.hhs.gov/surgeongeneral/priorities/youth-mental-health/social-media/index.html.

5. Woods HC, Scott H. #Sleepyteens: Social media use in adolescence is associated with poor sleep quality, anxiety, depression and low self-esteem. Journal of Adolescence. 2016 Aug;51:41–9. doi:10.1016/j.adolescence.2016.05.008.

6. O’Reilly M, Dogra N, Whiteman N, Hughes J, Eruyar S, Reilly P. Is social media bad for mental health and wellbeing? Exploring the perspectives of adolescents. Clinical Child Psychology and Psychiatry. 2018 Oct;23(4):601–13. doi:10.1177/1359104518775154.

7. McDougall MA, Walsh M, Wattier K, Knigge R, Miller L, Stevermer M, et al. The effect of social networking sites on the relationship between perceived social support and depression. Psychiatry Research. 2016 Dec;246:223–9. doi:10.1016/j.psychres.2016.09.018.

8. Lin LY, Sidani JE, Shensa A, Radovic A, Miller E, Colditz JB, et al. Association Between Social Media Use and Depression Among U.S. Young Adults. Depression and Anxiety. 2016 Apr;33(4):323–31. doi:10.1002/da.22466.

9. Pantic I, Damjanovic A, Todorovic J, Topalovic D, Bojovic-Jovic D, Ristic S, et al. Association between online social networking and depression in high school students: Behavioral physiology viewpoint. Psychiatria Danubina. 2012;24(1):90–3.

10. Griffiths MD, Kuss DJ, Demetrovics Z. Social Networking Addiction: An Overview of Preliminary Findings. In: Behavioral Addictions: Criteria, Evidence, and Treatment. London, UK: Academic Press; 2014. p. 119–41. doi:10.1016/B978-0-12-407724-9.00006-9.

11. Vannucci A, Flannery KM, Ohannessian CM. Social media use and anxiety in emerging adults. Journal of Affective Disorders. 2017 Jan;207:163–6. doi:10.1016/j.jad.2016.08.040.

12. Sá Sd, Baião A, Marques H, Marques MdC, Reis MJ, Dias S, et al. The Influence of Smartphones on Adolescent Sleep: A Systematic Literature Review. Nursing Reports. 2023;13:612. doi:10.3390/nursrep13020054.

13. Dibben GO, Martin A, Shore CB, Johnstone A, McMellon C, Palmer V, et al. Adolescents’ interactive electronic device use, sleep and mental health: A systematic review of prospective studies. Journal of Sleep Research. 2023;32:e13899. doi:10.1111/jsr.13899.

14. Burnell K, Garrett SL, Nelson BW, Prinstein MJ, Telzer EH. Daily links between objective smartphone use and sleep among adolescents. Journal of Adolescence. 2024;96:1171–81. doi:10.1002/jad.12326.

15. Nagata JM, Cheng CM, Shim J, Kiss O, Ganson KT, Testa A, et al. Bedtime Screen Use Behaviors and Sleep Outcomes in Early Adolescents: A Prospective Cohort Study. Journal of Adolescent Health. 2024;75:650–5. doi:10.1016/j.jadohealth.2024.06.006.

16. Campisi J, Folan D, Diehl G, Kable T, Rademeyer C. Social media users have different experiences, motivations, and quality of life. Psychiatry Research. 2015 Aug;228(3):774–80. doi:10.1016/j.psychres.2015.04.008.

17. Nabi RL, Prestin A, So J. Facebook friends with (health) benefits? Exploring social network site use and perceptions of social support, stress, and well-being. Cyberpsychology, Behavior, and Social Networking. 2013 Oct;16(10):721–7. doi:10.1089/cyber.2012.0521.

18. Shannon H, Bush K, Villeneuve PJ, Hellemans KGC, Guimond S. Problematic Social Media Use in Adolescents and Young Adults: Systematic Review and Meta-analysis. JMIR Mental Health. 2022;9(4):e33450. doi:10.2196/33450.

19. Megret C. No connectivity, better connections: teenagers’ experiences of a phone-free summer camp in the United States. Journal of Adventure Education and Outdoor Learning. 2023 May;24(1):65–78. Available from: 10.1080/14729679.2023.2211180. doi:10.1080/14729679.2023.2211180.

20. Kuo FE, Taylor AF. A potential natural treatment for attention-deficit/hyperactivity disorder: Evidence from a national study. American Journal of Public Health. 2004;94(9):1580–6.

21. Razani N, Radhakrishna R, Erdmann C, Chen Y, Rutherford G, Petray M. Greenspace exposure and sleep: A systematic review. Environmental Research. 2019;178:108682.

22. Blum RW, Lai J, Martinez M, Jessee C. Adolescent connectedness: cornerstone for health and wellbeing. BMJ. 2022;379:e069213. doi:10.1136/bmj-2021-069213.

23. Birrell L, Werner-Seidler A, Davidson L, Andrews JL, Slade T. Social connection as a key target for youth mental health. Mental Health & Prevention. 2025;37:200395. doi:10.1016/j.mhp.2025.200395.

24. Lamash L, Fogel Y, Hen-Herbst L. Adolescents’ social interaction skills on social media versus in person and the correlations to well-being. Journal of Adolescence. 2023 Sep;96(3):501–511. Available from: 10.1002/jad.12244. doi:10.1002/jad.12244.

25. Kaplan S. The restorative benefits of nature: Toward an integrative framework. Journal of Environmental Psychology. 1995 Sep;15(3):169–182. Available from: 10.1016/0272-4944(95)90001-2. doi:10.1016/0272-4944(95)90001-2.

26. Ulrich RS, Simons RF, Losito BD, Fiorito E, Miles MA, Zelson M. Stress recovery during exposure to natural and urban environments. Journal of Environmental Psychology. 1991 Sep;11(3):201–230. Available from: 10.1016/S0272-4944(05)80184-7. doi:10.1016/s0272-4944(05)80184-7.

27. Oberle E, Ji XR, Alkawaja M, Molyneux TM, Kerai S, Thomson KC, et al. Connections matter: Adolescent social connectedness profiles and mental well-being over time. Journal of Adolescence. 2023 Sep;96(1):31–48. Available from: 10.1002/jad.12250. doi:10.1002/jad.12250.

28. Whittington A, Garst BA, Gagnon RJ, Baughman S. Living without boys: A retrospective analysis of the benefits and skills gained at all-female camps. Journal of Experiential Education. 2017;40(2):97–113. doi:10.1177/1053825917690129.

29. Sorenson J. The fundamental characteristics and unique outcomes of Christian summer camp experiences. Journal of Youth Development. 2018;13(1-2):183–200. doi:10.5195/jyd.2018.546.

30. Povilaitis V. Smartphone-free summer camp: Adolescent perspectives of a leisure context for social and emotional learning. World Leisure Journal. 2019;61:276–90. doi:10.1080/16078055.2019.1661104.

31. Uhls YT, Michikyan M, Morris J, Garcia D, Small GW, Zgourou E, et al. Five days at outdoor education camp without screens improves preteen skills with nonverbal emotion cues. Computers in Human Behavior. 2014;39:387–92. doi:10.1016/j.chb.2014.05.036.

32. Thiagarajan TC, Tapert SF, Maxwell B, Giese CA, Parameshwaran D, Newson JJ. Impact of a device-free summer camp on adolescent mental health and wellbeing. Mental Health & Prevention. 2026:200554. doi:10.1016/j.mhp.2026.200554.

33. Zhang Y, Tang Y, Wang X, Tan Y. The Effects of Natural Window Views in Classrooms on College Students’ Mood and Learning Efficiency. Buildings. 2024 May;14(6):1557. Available from: 10.3390/buildings14061557. doi:10.3390/buildings14061557.

34. Al-barrak L, Kanjo E, Younis EMG. NeuroPlace: Categorizing urban places according to mental states. PLOS ONE. 2017 Sep;12(9):e0183890. Available from: 10.1371/journal.pone.0183890. doi:10.1371/journal.pone.0183890.

35. Luo Y, et al. Exposure to Natural Environments Alters Brain Activity: Evidence From EEG. Journal of Environmental Psychology. 2023;88:102010. doi:10.1016/j.jenvp.2023.102010.

36. Bailey AW, Kang HK. Walking and Sitting Outdoors: Which Is Better for Cognitive Performance and Mental States? International Journal of Environmental Research and Public Health. 2022 Dec;19(24):16638. Available from: 10.3390/ijerph192416638. doi:10.3390/ijerph192416638.

37. Jin Y, Yu Z, Yang G, Yao X, Hu M, Remme RP, et al. Quantifying physiological health efficiency and benefit threshold of greenspace exposure in typical urban landscapes. Environmental Pollution. 2024 Dec;362:124726. Available from: 10.1016/j.envpol.2024.124726. doi:10.1016/j.envpol.2024.124726.

38. Kang M, Kim S, Lee J. Pilot Study on the Physio-psychological Effects of Botanical Gardens on the Prefrontal Cortex Activity in an Adult Male Group. Journal of People, Plants, and Environment. 2022 Aug;25(4):413–423. Available from: 10.11628/ksppe.2022.25.4.413. doi:10.11628/ksppe.2022.25.4.413.

39. Youn C, Chung L, Kang M, Kim S, Choi H, Lee J. Effects of Green Walls on Prefrontal Cerebral Hemodynamics in Hospital Workers. Journal of People, Plants, and Environment. 2022 Dec;25(6):717–728. Available from: 10.11628/ksppe.2022.25.6.717. doi:10.11628/ksppe.2022.25.6.717.

40. Belkacem AN, Saetia S, Zintus-art K, Shin D, Kambara H, Yoshimura N, et al. Real-Time Control of a Video Game Using Eye Movements and Two Temporal EEG Sensors. Computational Intelligence and Neuroscience. 2015;2015:1–10. Available from: 10.1155/2015/653639. doi:10.1155/2015/653639.

41. Jijun T, Peng Z, Ran X, Lei D. The portable P300 dialing system based on tablet and Emotiv Epoc headset. In: 2015 37th Annual International Conference of the IEEE Engineering in Medicine and Biology Society (EMBC). IEEE; 2015. p. 566–569. Available from: 10.1109/EMBC.2015.7318425. doi:10.1109/embc.2015.7318425.

42. Vanhollebeke G, De Smet S, De Raedt R, Baeken C, van Mierlo P, Vanderhasselt MA. The neural correlates of psychosocial stress: A systematic review and meta-analysis of spectral analysis EEG studies. Neurobiology of Stress. 2022;18:100452. doi:10.1016/j.ynstr.2022.100452.

43. Newson JJ, Thiagarajan TC. EEG frequency bands in psychiatric disorders: A review of resting state studies. Frontiers in Human Neuroscience. 2019;12:521. doi:10.3389/fnhum.2018.00521.

44. Li Y, Tong S, Liu D, Gai Y, Wang X, Wang J, et al. Abnormal EEG complexity in patients with schizophrenia and depression. Clinical Neurophysiology. 2008;119(6):1232–41. doi:10.1016/j.clinph.2008.01.104.

45. Linkenkaer-Hansen K, Monto S, Rytsälä H, Suominen K, Isomets̈ä E, Kähkönen S. Breakdown of long-range temporal correlations in theta oscillations in patients with major depressive disorder. Journal of Neuroscience. 2005;25(44):10131–7. doi:10.1523/JNEUROSCI.3244-05.2005.

46. Robertson MM, Furlong S, Voytek B, Donoghue T, Boettiger CA, Sheridan MA. EEG power spectral slope differs by ADHD status and stimulant medication exposure in early childhood. Journal of Neurophysiology. 2019;122(6):2427–37. doi:10.1152/jn.00388.2019.

47. Gramfort A, Luessi M, Larson E, Engemann DA, Strohmeier D, Brodbeck C, et al. MEG and EEG Data Analysis with MNE-Python. Frontiers in Neuroscience. 2013;7(267):1–13. doi:10.3389/fnins.2013.00267.

48. Jas M, Engemann DA, Bekhti Y, Raimondo F, Gramfort A. Autoreject: Automated artifact rejection for MEG and EEG data. NeuroImage. 2017 Oct;159:417–429. Available from: 10.1016/j.neuroimage.2017.06.030. doi:10.1016/j.neuroimage.2017.06.030.

49. Donoghue T, Haller M, Peterson EJ, Varma P, Sebastian P, Gao R, et al. Parameterizing neural power spectra into periodic and aperiodic components. Nat Neurosci. 2020 Dec;23(12):1655–65. Number: 12 Publisher: Nature Publishing Group. Available from: https://www.nature.com/articles/s41593-020-00744-x. doi:10.1038/s41593-020-00744-x.

50. Gao R, Peterson EJ, Voytek B. Inferring synaptic excitation/inhibition balance from field potentials. Neuroimage. 2017 Sep;158:70–8. doi:10.1016/j.neuroimage.2017.06.078.

51. Lempel A, Ziv J. On the Complexity of Finite Sequences. IEEE Transactions on Information Theory. 1976 Jan;22(1):75–81. Available from: 10.1109/TIT.1976.1055501. doi:10.1109/tit.1976.1055501.

52. Fernández A, López-Ibor MI, Turrero A, Santos JM, Morón MD, Hornero R, et al. Lempel–Ziv complexity in schizophrenia: A MEG study. Clinical Neurophysiology. 2011 Nov;122(11):2227–2235. Available from: 10.1016/j.clinph.2011.04.011. doi:10.1016/j.clinph.2011.04.011.

53. Zhang Y, Hao J, Zhou C, Chang K. Normalized Lempel-Ziv complexity and its application in bio-sequence analysis. Journal of Mathematical Chemistry. 2008 Dec;46(4):1203–1212. Available from: 10.1007/s10910-008-9512-2. doi:10.1007/s10910-008-9512-2.

54. Vallat R. raphaelvallat/antropy: AntroPy, a Python toolbox for entropy and complexity of time-series. GitHub; 2024. https://github.com/raphaelvallat/antropy.

55. Linkenkaer-Hansen K, Nikouline VV, Palva JM, Ilmoniemi RJ. Long-range temporal correlations and scaling behavior in human brain oscillations. J Neurosci. 2001 Feb;21(4):1370–7. doi:10.1523/JNEUROSCI.21-04-01370.2001.

56. Thölke P, Arcand-Lavigne M, Lajnef T, Frenette S, Carrier J, Jerbi K. Caffeine induces age-dependent increases in brain complexity and criticality during sleep. Communications Biology. 2025 Apr;8(1). Available from: 10.1038/s42003-025-08090-z. doi:10.1038/s42003-025-08090-z.

57. Richman JS, Moorman JR. Physiological time-series analysis using approximate entropy and sample entropy. American Journal of Physiology-Heart and Circulatory Physiology. 2000 Jun;278(6):H2039–H2049. Available from: 10.1152/ajpheart.2000.278.6.H2039. doi:10.1152/ajpheart.2000.278.6.h2039.

58. Richman JS, Lake DE, Moorman JR. In: Sample Entropy. Elsevier; 2004. p. 172–184. Available from: 10.1016/S0076-6879(04)84011-4. doi:10.1016/s0076-6879(04)84011-4.

59. Makowski D, Pham T, Lau ZJ, Brammer J, Lespinasse F, Pham H, et al. NeuroKit2: A Python toolbox for neurophysiological signal processing. Behavior Research Methods. 2021;53(4):1689–96. doi:10.3758/s13428-020-01516-y.

60. Zhang A, Yang B, Huang L. Feature extraction of EEG signals using power spectral entropy. In: 2008 International Conference on BioMedical Engineering and Informatics. IEEE; 2008..

61. Newson JJ, Thiagarajan TC. Assessment of population well-being with the Mental Health Quotient (MHQ): Development and usability study. JMIR Mental Health. 2020;7:e17935. doi:10.2196/17935.

62. Newson JJ, Pastukh V, Thiagarajan TC. Assessment of population well-being with the Mental Health Quotient: Validation study. JMIR Mental Health. 2022;9:e34105. doi:10.2196/34105.

63. Newson JJ, Sukhoi O, Thiagarajan TC. MHQ: Constructing an aggregate metric of population mental wellbeing. Population Health Metrics. 2024;22:16. doi:10.1186/s12963-024-00336-y.

64. Bailey AW, Kang H. Nature Exposure and EEG Activity: A Systematic Review. International Journal of Environmental Research and Public Health. 2022;19(24):16638. doi:10.3390/ijerph192416638.

65. Jiang B, et al. The Psychophysiological Effects of Viewing Nature: EEG Evidence. Environment and Behavior. 2019;51(9-10):1079–103. doi:10.1177/1420326X19870578.

66. Parameshwaran D, Thiagarajan TC. Modernization, wealth and the emergence of strong alpha oscillations in the human EEG. bioRxiv. 2017 Apr. Available from: 10.1101/125898. doi:10.1101/125898.

67. Medel V, Irani M, Ossandón T, Boncompte G. Complexity and 1/f slope jointly reflect cortical states across different E/I balances. Scientific Reports. 2020;13:21700. doi:10.1038/s41598-023-48639-0.

68. Gao R, Peterson EJ, Voytek B. Inferring synaptic excitation/inhibition balance from field potentials. NeuroImage. 2017;158:70–8. doi:10.1016/j.neuroimage.2017.06.078.

69. Trakoshis S, et al. Intrinsic excitation–inhibition imbalance affects medial prefrontal cortex differently in autistic men versus women. eLife. 2020;9:e55684. doi:10.7554/eLife.55684.

70. Medel V, Irani M, Crossley N, Ossandón T, Boncompte G. Complexity and 1/f slope jointly reflect brain states. Scientific Reports. 2023 Dec;13(1). Available from: 10.1038/s41598-023-47316-0. doi:10.1038/s41598-023-47316-0.

71. Höhn C, Hahn MA, Lendner JD, Hoedlmoser K. Spectral Slope and Lempel–Ziv Complexity as Robust Markers of Brain States during Sleep and Wakefulness. eneuro. 2024 Mar;11(3):ENEURO.0259-23.2024. Available from: 10.1523/ENEURO.0259-23.2024. doi:10.1523/eneuro.0259-23.2024.

